# Derivation of the effective reproduction number ℛ for COVID-19 in relation to mobility restrictions and confinement

**DOI:** 10.1101/2020.04.06.20054320

**Authors:** Alex Arenas, Wesley Cota, Jesús Gómez-Gardeñes, Sergio Gómez, Clara Granell, Joan T. Matamalas, David Soriano-Paños, Benjamin Steinegger

## Abstract

The spread of COVID-19 is posing an unprecedented threat to health systems worldwide^1^. The fast propagation of the disease combined with the existence of covert contagions by asymptomatic individuals make the controlling of this disease particularly challenging. The key parameter to track the progression of the epidemics is the effective reproduction number ℛ, defined as the number of secondary infections generated by an infected individual^2^. The suppression of the epidemics is directly related to this value, and is attained when ℛ < 1. Here, we find an analytical expression for ℛ as a function of mobility restrictions and confinement measures, using an epidemic model tailored for COVID-19. This expression for ℛ is an extremely useful tool to design containment policies that are able to suppress the epidemics. We applied our epidemic model for the case of Spain, successfully forecasting both the observed incidence in each region and the overload of the health system. The expression for ℛ allowed us to determine the precise reduction of mobility *κ*_0_ needed to bend the curve of epidemic incidence, which turned out to be *κ*_0_ ∼ 0.7. This value, for the case of Spain, translates to a total lockdown with the exception of the mobility associated to essential services, a policy that was finally enforced on March 28.

## 1 Introduction

The past 31 December of 2019 an outbreak of a novel coronavirus, named SARS-CoV-2 and responsible for causing the COVID-19 disease, was reported in Hubei, China. As of 30 March of 2020, more than 178 countries/regions have reported cases, and community transmission of the virus inside those regions is growing unceasingly. The current pandemic is most likely one of the most serious challenges that our interconnected modern society will face, and requires for pre-emptive and coordinated actions to decelerate transmission globally. Such measures should give nations a precious time to prepare their health-care systems to be capable of withstanding the impact of COVID-19.

The confinement of populations, from regions to entire countries, remains the best non-pharmacological intervention once traceability of individual cases —only feasible under massive testing— is lost. However, the success of such measures is highly dependent on the precise time they are implemented, the extent of the mobility restrictions, and the total duration of the confinement. Normally, the decisions regarding such measures are made taking the number of reported cases as a proxy of the current disease spread status. However in many epidemics, and especially in the case of COVID-19, this information is an underestimation of the real prevalence in the population, due to the lack of widespread testing and the number of asymptomatic infections that hamper traceability of secondary cases.

Recently, we have observed how some countries, such as Spain or Italy, have failed in implementing the appropriate measures to stop the advance of the disease on time. Our model suggests this is due to the fact that the mobility reduction imposed was insufficient to bend the curve of incidence of COVID-19. According to the best estimates, the confinement measures enforced in Spain on March 14 achieved only a mobility reduction of 60 ± 5%^3, 4^ during the first week, a quantity that, according to our model, falls short to drive the system below the targeted ℛ < 1.

This example illustrates the difficult task of making accurate early evaluations of containment measures. It is observed that the reduction of mobility due to the confinement of the population has a highly nonlinear effect on the reduction of the epidemic. This makes the task of policy makers notoriously difficult, as they need to design realistic, implementable, confinement measures that reduce the impact of the epidemic sufficiently within the shortest time frame possible. If the interventions planned are too weak, the epidemic will not cease and the surge capacity of healthcare systems will be surpassed, leaving many individuals requiring hospitalization out of the attention they need. Furthermore, once policy makers realize the insufficiency of the interventions put in place, the epidemic may have spread up to a point such that only the most severe interventions will be of real use. On the other side, implementing very strict confinement measures from an early stage would give governments a much needed time to prepare against the attack of the disease, but would most likely take a huge toll in the economy, and consequently, in the population.

Finding the right balance in the intervention measures is a daunting, but essential task for successfully weathering the storm caused by this and any other epidemic in the future. To succeed in this endeavor, predicting the impact of any envisaged actions is crucial. We need to know the exact relationship between the reduction of mobility and confinement measures and its impact on the reproductive ratio ℛ of the epidemics, as this would allow policy makers to implement just the right amount of mobility restrictions needed to minimize the impact of the epidemic.

In this paper we aim to solve this problem. We analytically relate the ℛ of the evolution of COVID-19, in a demographically heterogeneous structured population, to the mobility patterns and the permeability of households under confinement. We derive a mathematical expression that allows us to anticipate the result of mobility restrictions and confinement on the effective reproduction number of the spreading of COVID-19 in any specific region. The dependence of ℛ on the average mobility is nonlinear, and presents a sudden transition at ℛ = 1. This transition separates a regime in which the mobility restrictions smoothly slow down the number of cases —the effect known as *flattening the curve*—, to a regime in which these restrictions sharply decelerate the number of cases —which we refer to as *bending the curve*—. The difference in the outcome of these two scenarios is very pronounced. First, the curve bending strategy provokes a drastic reduction in the infection speed, attaining a lower attack rate than the flattening strategy. Secondly, under the bending strategy, the mobility restriction and confinement measures have to be enforced for a shorter amount of time. This makes the bending strategy the most appropriate way of handling the confinement measures. Nevertheless, the question remains, given an interconnected, structured population, what is the amount of mobility reduction needed to drive the epidemic into the bending regime. The aforementioned analytical relationship between ℛ and the mobility restrictions precisely allows us to answer this question for any region under the attack of COVID-19 for which demographic and mobility data is available.

## 2 Results

In order to explicitly relate the reduction of mobility and confinement measures with the reproductive ratio ℛ of the epidemics, we first develop a mathematical model, in the line of previous approaches^5–11^, to predict the epidemic prevalence of COVID-19 throughout a territory. We develop an age-stratified metapopulation compartmental model that encapsulates the particularities of the spreading of COVID-19 regarding (i) its transmission among individuals, (ii) the specificities of certain demographic groups with respect to the impact of COVID-19, and (iii) the human mobility patterns inside and among regions. The full dynamics of the epidemics is formalized in terms of a Microscopic Markov Chain Approach (MMCA)^12–14^. We refer the reader to the Methods section for a full description of the model and to the Supplementary Note 1 for the complete analysis of the Markovian equations. This model has been used to assess the necessity for total lockdown in Spain, finally enforced on March 30th^15^, for it allows a direct assessment of mobility restrictions and confinement policies.

The effective reproduction number ℛ(*t*) is defined as the number of secondary cases that an individual, becoming infectious at time *t*, will produce over time. To achieve the computation of this quantity, and understand its dependencies, we present an incremental rationale from the most rough approximation to the most accurate formula.

Let us consider, first, an scenario (in physics know as mean-field) in which an infected subject *i* makes ⟨*k*⟩ contacts each time step. Assuming an infection probability *β*, the expected number of individuals infected by *i* at each time step is *β*⟨*k*⟩*ρ*_*S*_, where *ρ*_*S*_ is the fraction of susceptible individuals on the population. We can estimate how many individuals have been infected by subject *i* over time as:

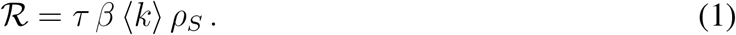

This mean-field approach, reveals the main dependencies of ℛ on the variables of the epidemic propagation. However, this approximation obviates the mobility of individuals and the age strata, as well as the temporal evolution of the variables. This can be largely improved by leveraging the information of the current epidemic model.

Let us define 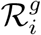, (the reproduction number of patch *i* and age group *g*) as the number of secondary cases produced by an infected individual belonging to patch *i* and age group *g*^16^. Mathematically, 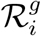 is expressed as:

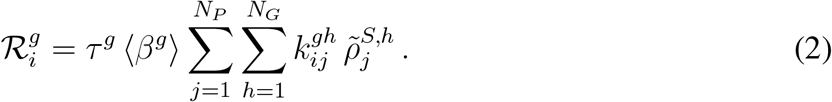

In the above expression the following quantities interplay: the duration of the infectious period, *τ* ; the average infection probability during the infectious period, ⟨*β*^*g*^⟩ per age group *g*; the average number of daily contacts an individual belonging to patch *i* and age group *g* makes with individuals in patch *j* belonging to age group 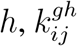; and the fraction of susceptible individuals present in patch *j* belonging to age group 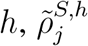. In summary, the structure of 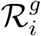 resembles the mean field expression presented in Eq. 1 but takes explicitly into account the mobility of individuals (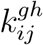 and 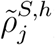). See Methods and Supplementary Note 5 for further details.

The last step to develop a formula for the evolution of ℛ consists in including the temporal dependence on the variables. Both, the average number of contacts 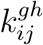 as well as the pool of susceptible individuals 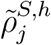 vary in time. Furthermore, we need to take into account the transitions between, and out of, the infectious compartments of the epidemic model in the evolution of an infected individual. Incorporating these elements we obtain:

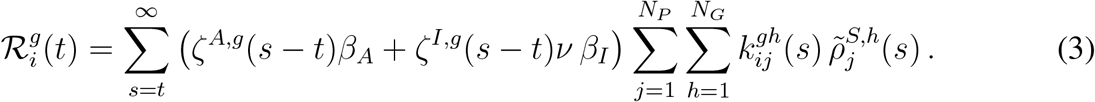

Functions *ζ*^*A*,*g*^ and *ζ*^*I*,*g*^ account for the probability for an individual, which becomes asymptomatically infectious at time *t*, to be in the asymptomatic and infected state, respectively, at each posterior time step (see Methods for details). We suppose different values of the infection probability for the asymptomatic (*β*_*A*_) and symptomatic (*β*_*I*_) individuals, and we add a factor *ν* to account for the possible isolation of symptomatic individuals by themselves.

Once the effective reproduction number is computed for each patch, the value of ℛ(*t*) is the weighted average over all values 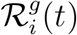 taking into account the distribution of the total population *N* across patches and age groups, 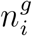, the final analytical expression for the evolution of the effective reproduction number ℛ(*t*) is:

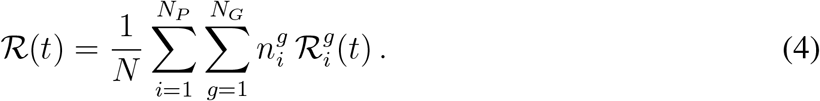

This expression corresponds to the main result of our study. It allows for evaluating the evolution of the effective reproductive number throughout an epidemic, taking into account the epidemiological characteristics of the disease along with the social, demographic and mobility patterns that characterize the affected population.

It is precisely the inclusion of social, demographic and mobility data in the expression of ℛ(*t*) what makes this expression a fundamental tool to evaluate and anticipate the effects of confinement and the reduction of mobility. Containment measures are implemented in the model in the spirit of Maier et al.^17^. In this line, the containment measures confine a fraction *κ*_0_ of the population at a given time *t*. The confinement leads to a reduction in the number of daily contacts to the household size and mobility. Furthermore, the confined population is subtracted from the pool of susceptible individuals but corrected accordingly to the permeability of the households *ϕ* (understood as the probability of members of a household to skip confinement for purchase of essential goods). Including these containment measures in the dynamical equations of the proposed epidemic model is straightforward, see Supplementary Note 2.

We first show that our model is able to capture the effects of implementing confinement measures by using demographic and mobility data for Spain (see Methods section). Once initialized the equations with the initial infectious seeds with the first reported cases up to March 3, we let the model evolve in the free-mobility regime until March 14, when the first mobility restrictions enter into play in Spain and, consequently, confinement is imposed in the model. Figure 1 compares the predictions of the model for the daily incidence and the number of cases requiring Intensive Care Unit (ICU) assistance with the official data reported by Spain Health Ministry. There it becomes clear that the model, not only qualitatively but also quantitatively, reproduces the impact of restraining human mobility and promoting social distancing on hampering the spread of COVID-19. In the Supplementary Figure 2 we also show the same comparison for daily reported cases at the level of autonomous regions in Spain.

**Figure 1.**
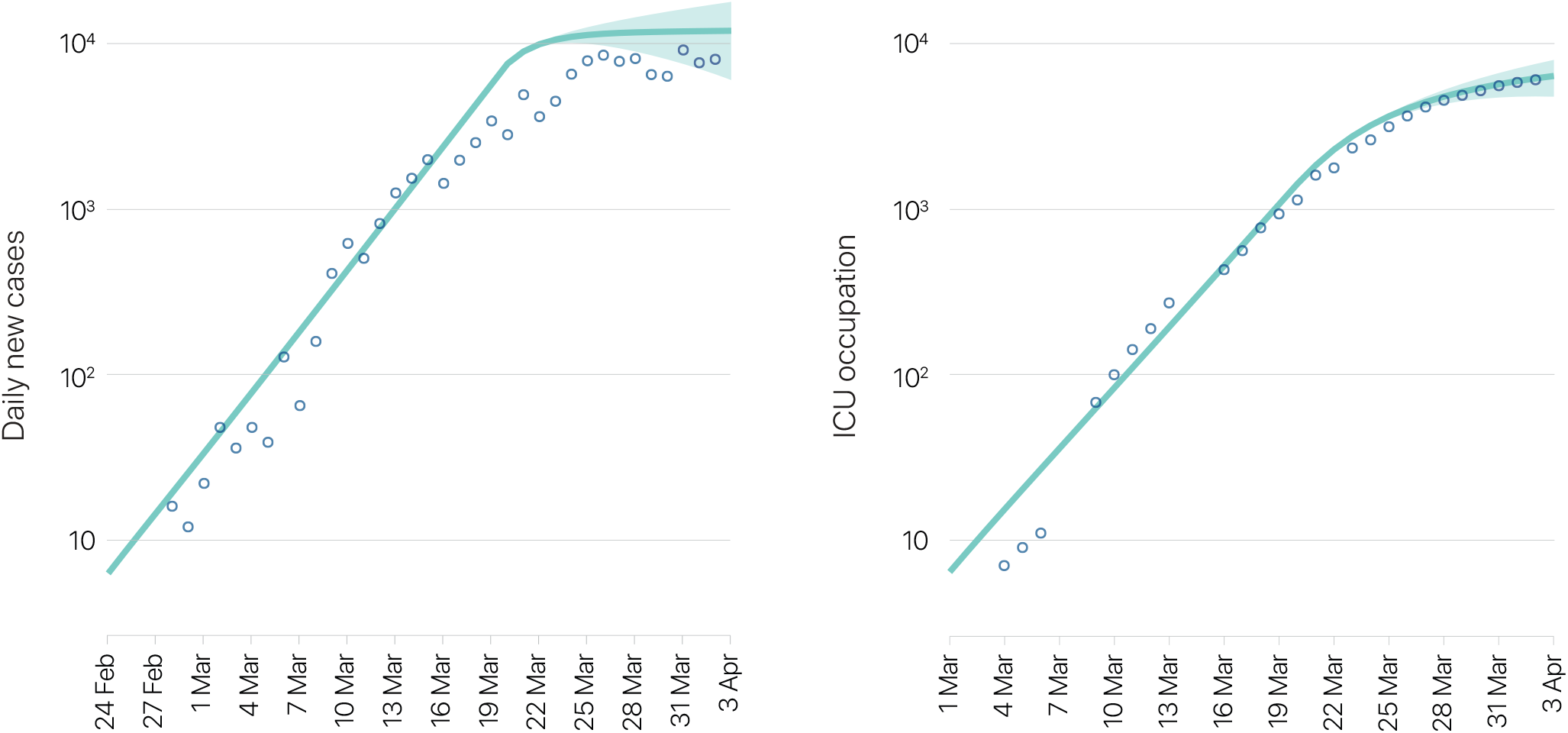
Prediction of the number of cases and the number of patients requiring ICU assistance compared to reported cases for Spain. Left: The solid line is the result of the epidemic model, aggregated by ages, for the number of individuals inside compartments (H+R+D) that corresponds to the expected number of daily reported cases, and dots correspond to real cases reported (mean absolute error: 1974.14). Right: Inpatients in intensive care units (ICU) in Spain. The solid dark line represents the number of patients predicted in compartment H by the model and dots correspond to real ICU occupation (mean absolute error: 181.83). The containment strategy has been applied on March 14 on both figures averaging results from *κ*_0_ in the [0.6, 0.8] interval and *ϕ* in on a range of [0.2, 0.4]. The red area covers an interval of ±2 standard deviations.

Once validated the model, we study how enforcing mobility restriction and social distancing can drastically change the course of the epidemics. In the left panel of Figure 2, we show the number of new cases per day for different levels of confinement, *κ*_0_. The real value of *κ*_0_ has been assessed from official data on the use of public transportation, cars entrance and exits in cities, and pedestrians flows tracked through cellphones. The confinement is applied at time *t*_*c*_ corresponding to March 14. It is observed that, as confinement is applied to a small fraction of the population (*κ*_0_ > 0), the curve for the number of new cases per day starts to broaden and the maximum shifts forward in time. Such a mitigation strategy is known as –flattening the curve–, in which containment delays and lowers the incidence peak. The consequence of the flattening scenario is that the impact over health systems is ameliorated at the expense of a larger epidemic period. However, as confinement is increased (higher *κ*_0_), there is a dramatic change for *κ*_0_ between 0.6 − 0.8 in the behavior of the epidemic curve. In particular, for large enough values of *κ*_0_ the epidemic curve reaches its maximum soon after containment is put in place, achieving a completely new scenario, referred to as –bending the curve–, in which prevalence decrease steadily and the epidemic wave is shortened. The two regimes are separated by the condition ℛ(*t*_*c*_) translating into a critical value of confinement, 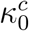.

**Figure 2.**
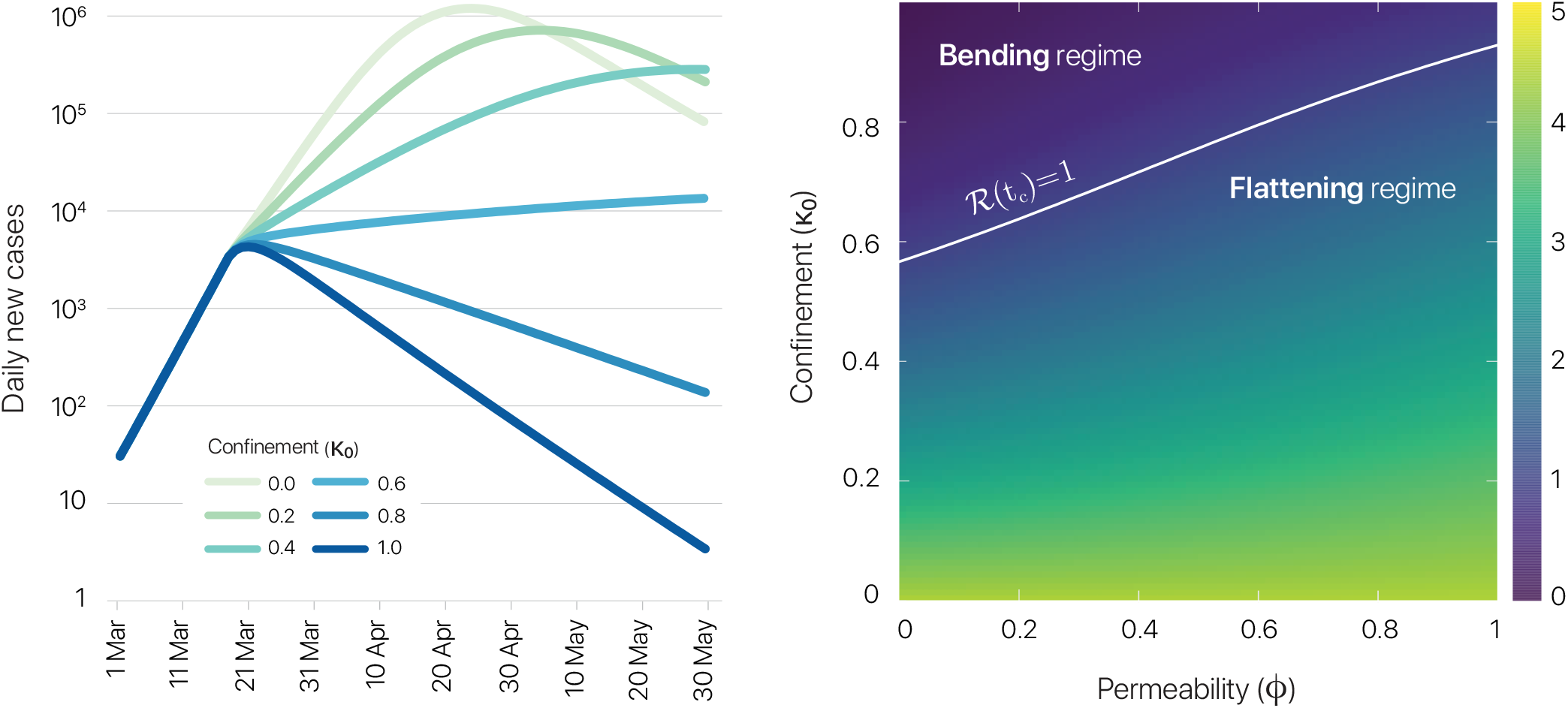
Relationship between ℛ and the mobility restrictions and confinement. Left: Temporal evolution of the number of daily new reported cases in Spain (see text for further details) as a function of the confinement *κ*_0_ (color code). The permeability has been set to *ϕ* = 0.2. Right: Effective reproduction number when mobility restrictions are imposed ℛ(*t*_*c*_) (color code) as a function of the confinement *κ*_0_ and the household permeability *ϕ*. The white line denotes the condition ℛ(*t*_*c*_) = 1 separating the different regimes for which enforcing the confinement leads to the flattening or bending of the epidemic curve respectively. See Supplementary Note 8 and Supplementary Figure 4 for an analysis of the sensitivity of the results.

The two regimes are separated by the condition ℛ(*t*_*c*_) = 1. The transition between the two regimes is indicated by a white solid line in the right panel of Figure 2, which shows the effective reproduction number as function of the confinement, *κ*_0_, and the permeability of the households, *ϕ*. The bigger the permeability of the households, i.e. the softer the confinement, the higher the fraction of the population that needs to be confined in order to bend the curve.

To illustrate the sudden change in ℛ(*t*) caused by the implementation of a confinement triggering a subcritical regime 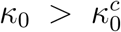, in Figure 3 we show the evolution of ℛ(*t*) in the case of Spain when confinement is applied on March 14, 2020. Furthermore, we present the evolution of the effective reproduction number of adults (*g* = *M*) for each municipality in Spain, showing that they reach 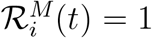 in different stages. This reveals that the decision to relax containment measures must take into account the entire set of municipalities and not be based exclusively on the effective reproduction number of the country. In particular, relaxing the confinement before every single age group *g* within each patch *i* fulfills the condition 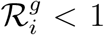 can lead to secondary outbreaks driven by those most vulnerable areas. To further illustrate this phenomenon, we show in Figure 3 the spatial distribution of the effective reproduction number of the adult population 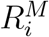 over the country before and after confinement is implemented. See also Supplementary Figure 3 for the distribution of 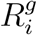 among different patches for two different temporal snapshots, before and after confinement.

**Figure 3.**
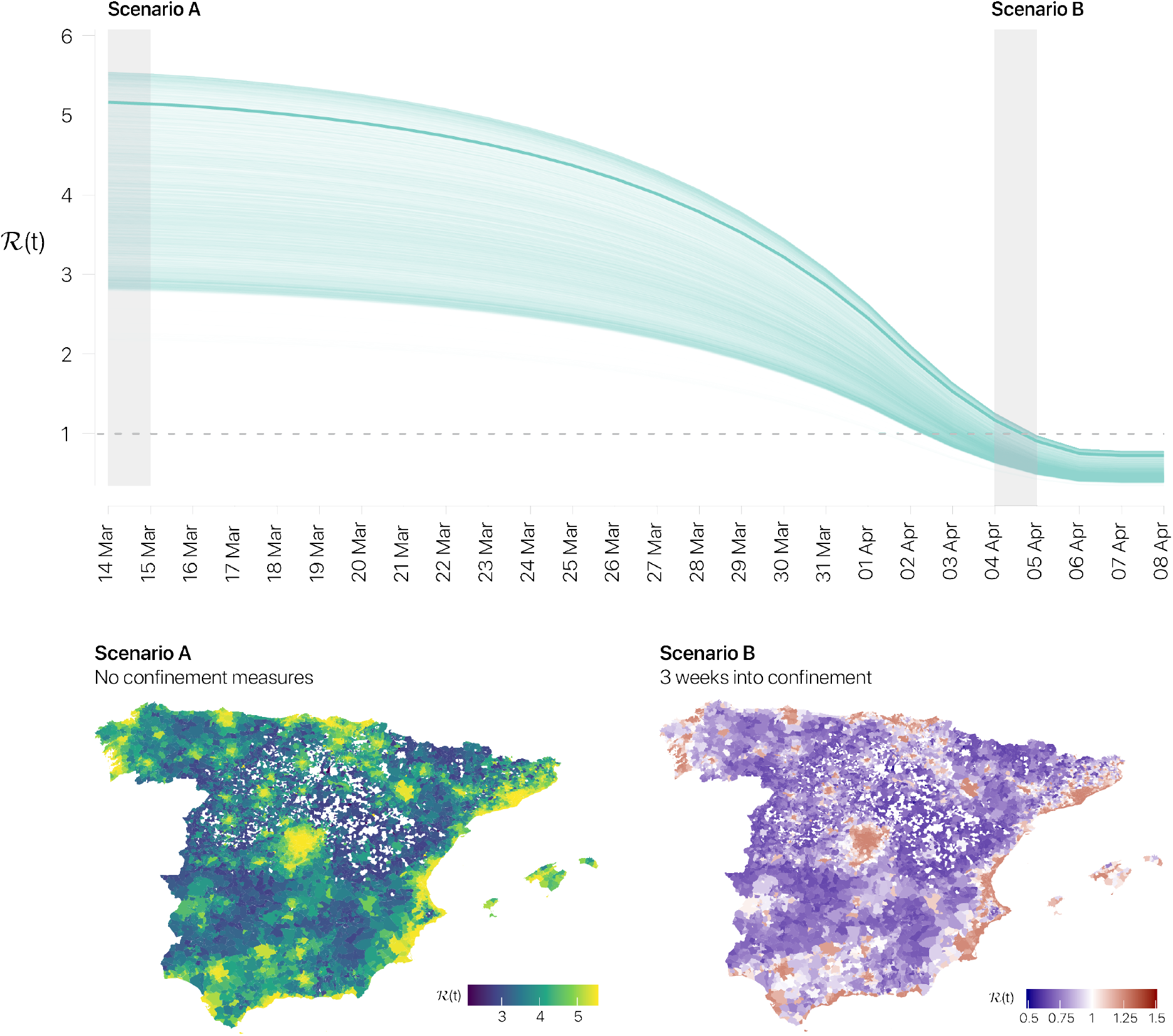
Prediction of the evolution of ℛ. Top: Temporal evolution of the effective reproduction number associated to each age group *g* within each municipality *i*. The solid dark line depicts the global effective reproductive number computed according to Eq. (4). The values of ℛ in the plot are shifted in time to account for the lag between the definition of ℛ and the one usually estimated from data, see EpiEstim^20^, plus the lag in the reporting detected cases, a total of 20 days. Bottom: Spatial distribution of the effective reproduction number of adult population, 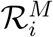, before and after confinement policy is promoted. For both panels, confinement is implemented on 14 March by setting (*κ*_0_, *ϕ*) = (0.70, 0.20). These values correspond to the mobility officially reported by authorities in Spain.

## 3 Conclusions

As the COVID-19 pandemic affects more and more countries and threatens to overload the critical capacity of health systems, the absence of a SARS-CoV-2 vaccine requires forceful non-pharmacological containment measures by governments. Among these measures, the restriction of mobility and social distancing have been implemented by many countries in an attempt to reduce the impact on health systems and save the time necessary to try more efficient treatments against this emerging pathogen. However, these measures have come up short in Spain to cut the progression of COVID-19, due to covert infections that characterize its silent and rapid transmission beyond areas with a high number of cases detected.

To mount the strategy against SARS-CoV-2 and anticipate its trajectory, it is necessary, in addition to correctly modeling its epidemiological characteristics, to take into account as closely as possible the influence of the fluidity of social contacts that it uses to spread. Here we have shown that it is possible to construct the expression of the effective reproduction number ℛ(*t*) capturing both the epidemiological characteristics of COVID-19 and those social patterns that facilitate its expansion. This expression enables an accurate evaluation of the spreading potential of SARS-CoV-2 on a given population and, moreover, the assessment of non-pharmacological interventions.

Focusing to the current outbreak in Spain, we have analyzed the effects of different degrees of mobility restrictions *κ*_0_, using the expression for ℛ(*t*) evaluated at the time *t*_*c*_ when containment is applied. This way, we can accurately determine the effects on the slowdown and suppression of the epidemic caused by different degrees of confinement. Calculating the value ℛ(*t*_*c*_) we observe a transition as the set of confined inhabitants and the consequent social distancing increase. This transition defines a critical confinement, 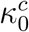, that separates a supercritical scenario, in which ℛ(*t*_*c*_) > 1 giving rise to a flattening of the curve, to another subcritical one, ℛ(*t*_*c*_) < 1, in which the change in the social structure is so profound that bends the epidemic curve making it impossible for the virus to spread.

The exact critical confinement separating the supercritical and subcritical regimes is highly dependent on the underlying social structure and the intrinsic mobility patterns of each population. Furthermore, it depends on the time it is applied, since it depends on the available pool of susceptible agents that can be culled out from the system. The generality of the expression ℛ(*t*) provided here makes it possible to apply it to any population, paving the way for implementing timely and well-founded epidemiological and socially based non-pharmacological responses.

## Data Availability

Code/data is available upon request.

## Methods

### Markovian epidemic model

The disease dynamics is described making use of variables 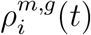 which represent the probabilities that individuals of age group *g* in region *i* are in state *m* at time *t*, where *m* take the values *S*, *E*, *A*, *I*, *H*, *D* and *R*, and the age group *g* is either *Y*, *M* or *O*. The equations of the model read as:

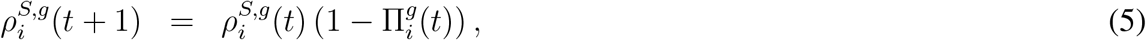

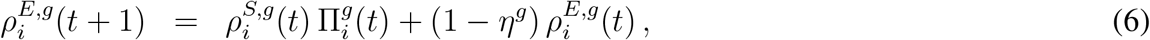

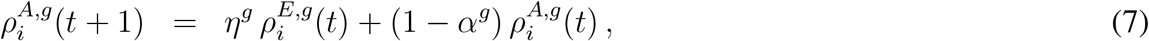

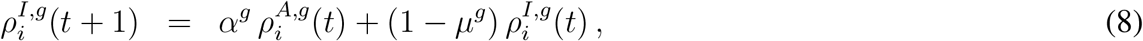

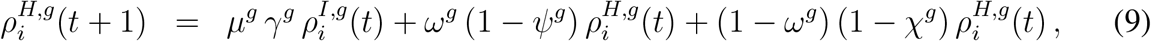

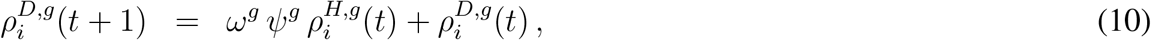

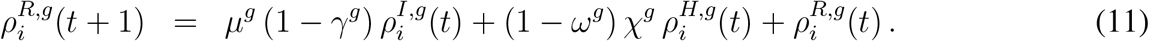

These equations correspond to a discrete-time dynamics, where each time-step represents a day.

The rationale of the model is the following. Susceptible individuals become exposed by contacts with asymptomatic and infected subjects, with a probability 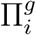. Exposed individuals turn into asymptomatic at a certain rate *η*^*g*^, which in turn become infected at a rate *α*^*g*^. Once infected, two paths emerge, which are reached at an escape rate *µ*^*g*^. The first option is requiring hospitalization in an ICU, with a certain probability *γ*^*g*^; otherwise, the individuals become recovered. While being at ICU, individuals have a death probability *ω*^*g*^, which is reached at a rate *ψ*^*g*^. Finally, ICUs discharge at a rate *χ*^*g*^, leading to the recovered compartment. See Supplementary Table 1 for a summary of the parameters of the model, and their values to simulate the spreading of COVID-19 in Spain.

The value of 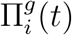 encodes the probability that a susceptible individual belonging to age group *g* and region *i* contracts the disease. Under the model assumptions, this probability is given by:

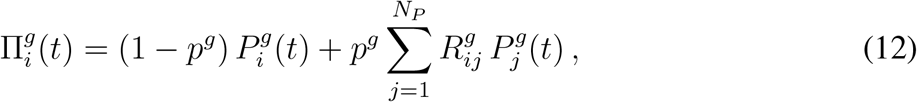

where *p*^*g*^ denotes the degree of mobility of individuals within age group *g*, and 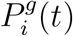 denotes the probability that those individuals get infected by the pathogen inside region *i*. This way, the first term in the r.h.s. of Eq. (12) denotes the probability of contracting the disease inside the residence patch, whereas the second term contains those contagions taking place in any of the neighboring areas. The expression for 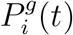 is quite involved, since it takes into account the infection probabilities, the average number of contacts of the individuals, the population density in each region, the probabilities of contacts between the different age strata, and the isolation of infected individuals; see Supplementary Note 1 for the full description of the model.

### Dynamical approximation of the effective reproduction number

In the expression of the effective reproduction number 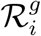 for region *i* and age group *g*, Eq. (2), we have separated the contributions corresponding to the average number of contacts made at patch *j* with people of age strata 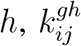, and the probability of finding a susceptible individual in patch 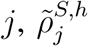. We need to relate them with all the parameters and variables of the model.

In our model, the number of contacts takes into account the following factors: the average number of contacts of people of age group *g, k*^*g*^ ; a nonlinear increasing function of the density of the population in the patch, *z*^*g*^*f* (*ñ*_*j*_*/s*_*j*_), where *s*_*j*_ is the area of the patch, *ñ*_*j*_ is the population at the patch while commuting takes place, and *z*^*g*^ is a normalization factor for the density across patches; the probabilities of contact according to age, *C*^*gh*^; and the probability of moving from patch *i* to patch *j, M*_*ij*_. With these ingredients, the expression for the number of contacts is:

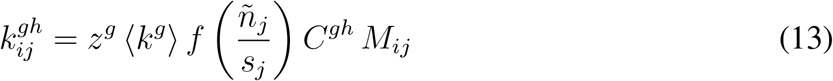

The probability of finding a susceptible individual of age *h* in patch *j* is simply given by the fraction of susceptible people coming from the rest of the patches:

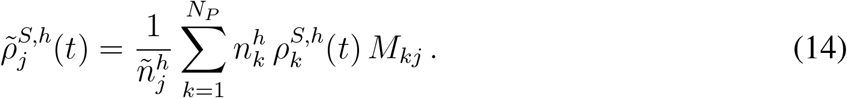

Finally, the expected infectious period is given by

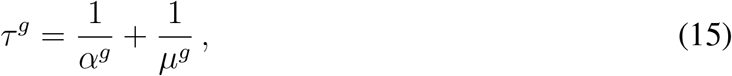

and the average infection probability by

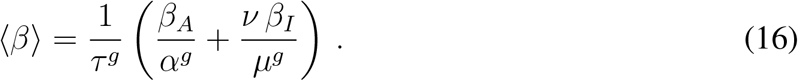

See Supplementary Note 5 for full details.

### Dependence on time of the effective reproduction number

In Eq. (2), we assume that the fraction of susceptible individuals as well as the average number of contacts is constant during the infectious period. However, the fraction of susceptible individuals obviously changes during that time. More importantly, as containment measures are put in place, the number of contacts varies as well. To account for the temporal variability of these quantities, we calculate the contributions to 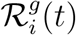 of the infections produced since the individual became asymptomatically infected at time *t*. Since the infectious period is divided in two phases, one asymptomatic with infectious rate *α*^*g*^, and another symptomatic with infectious rate *µ*^*g*^, what we have is a coupled Poisson process. The evolution of the probabilities to be asymptomatic, *ζ*^*A*,*g*^, or symptomatic, *ζ*^*I*,*g*^, are given by:

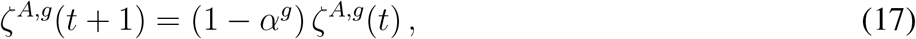

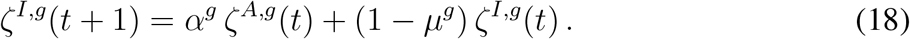

Since the seed is initially placed as asymptomatic, the initial conditions are given by *ζ*^*A*,*g*^(0) = 1 and *ζ*^*I*,*g*^(0) = 0. Accordingly, solving explicitly the recurrent equations, we find:

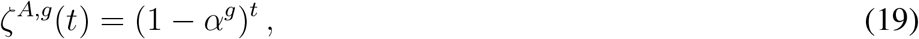

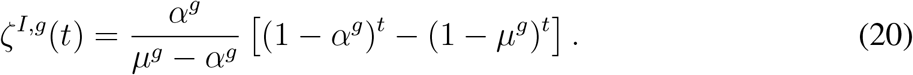

Incorporating these probabilities into Eq. (2) we obtain Eq. (3).

### Data from Spain

Regarding the population structure in Spain, we have obtained the population distribution, population pyramid, daily population flows and average household size at the municipality level from Instituto Nacional de Estadística ^18^ whereas the age-specific contact matrices have been extracted from ^19^. The first imported cases in Spain from January 31 to March 3 were used as the initial conditions of the dynamics. See Supplementary Note 6 and Supplementary Table 1 for the description and selection of the parameters of the model, and Supplementary Note 7 for more details on the calibration and validation of the model for the prediction of the evolution of the spreading of COVID-19 in Spain.

#### Acknowledgments

We thank Gourab Ghoshal and Silvio Ferreira for useful discussions. We also thank Matteo Rini for his suggestions. A.A., B.S. and S.G. acknowledge financial support from Spanish MINECO (grant PGC2018-094754-B-C21), Generalitat de Catalunya (grant No. 2017SGR-896), and Universitat Rovira i Virgili (grant No. 2018PFR-URV-B2-41). A.A. also acknowledge support from Generalitat de Catalunya ICREA Academia, and the James S. McDonnell Foundation grant #220020325. J.G.G. and D.S.P. acknowledges financial support from MINECO (projects FIS2015-71582-C2 and FIS2017-87519-P) and from the Departamento de Industria e Innovacioń del Gobierno de Aragoń y Fondo Social Europeo (FENOL group E-19). C.G. acknowledges financial support from Juan de la Cierva-Formacioń (Ministerio de Ciencia, Innovacioń y Universidades). B.S. acknowledges financial support from the European Union’s Horizon 2020 research and innovation program under the Marie Sklodowska-Curie grant agreement No. 713679 and from the Universitat Rovira i Virgili (URV). W.C. acknowledges financial support from the Coordenação de Aperfeiçoamento de Pessoal de N ível Superior, Brasil (CAPES), Finance Code 001.

## Contributions

All of the authors wrote the paper and contributed equally to the production of the manuscript.

## Competing financial interests

The authors declare no competing financial interests.

## Additional information

Supplementary Information is available for this paper.

Correspondence and requests for materials should be addressed to Alex Arenas and Jesús Gómez-Gardeñes.

## Derivation of the effective reproduction number ℛ for

### Supplementary Information

**Supplementary Figure 1:**
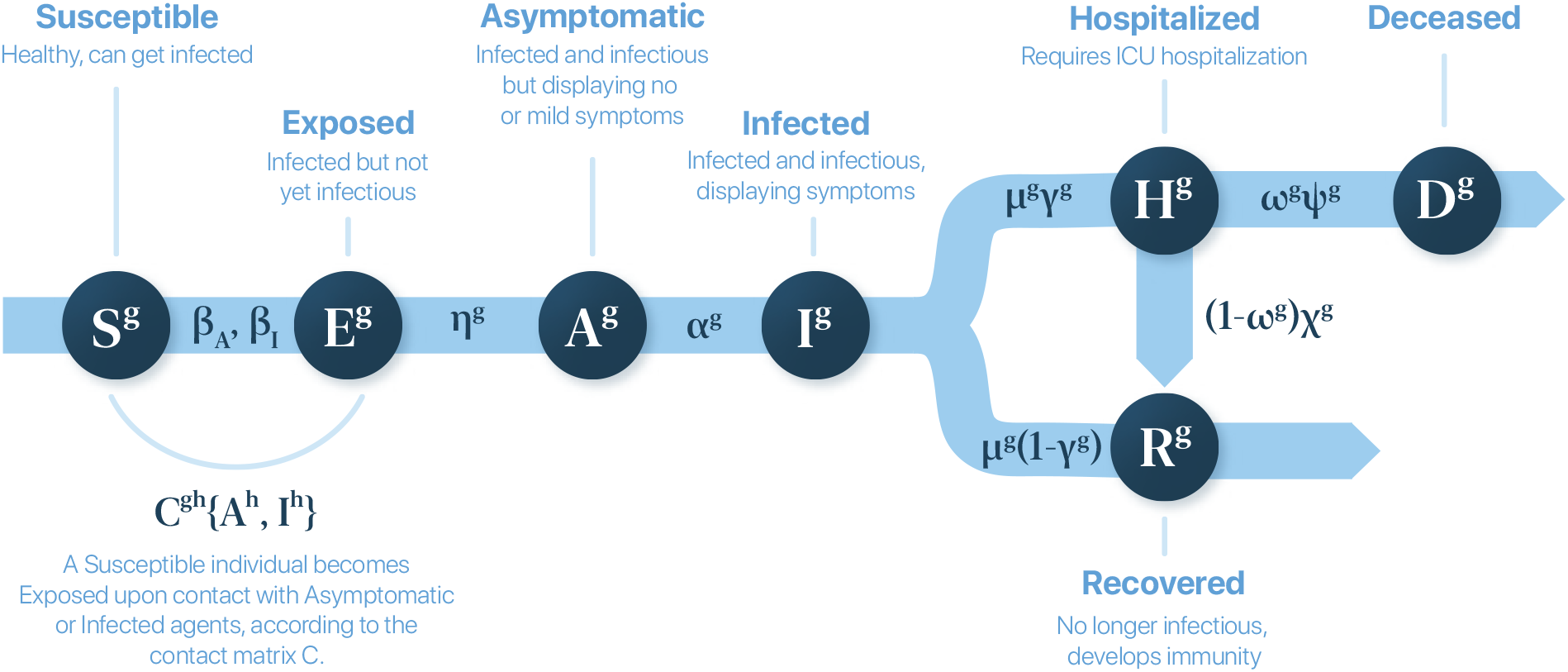
Compartmental epidemic model proposed in this study. The acronyms are susceptible (*S*^*g*^), exposed (*E*^*g*^), asymptomatic infectious (*A*^*g*^), infected (*I*^*g*^), hospitalized to ICU (*H*^*g*^), dead (*D*^*g*^), and recovered (*R*^*g*^), where *g* denotes the age stratum for all cases.

**Supplementary Figure 2:**
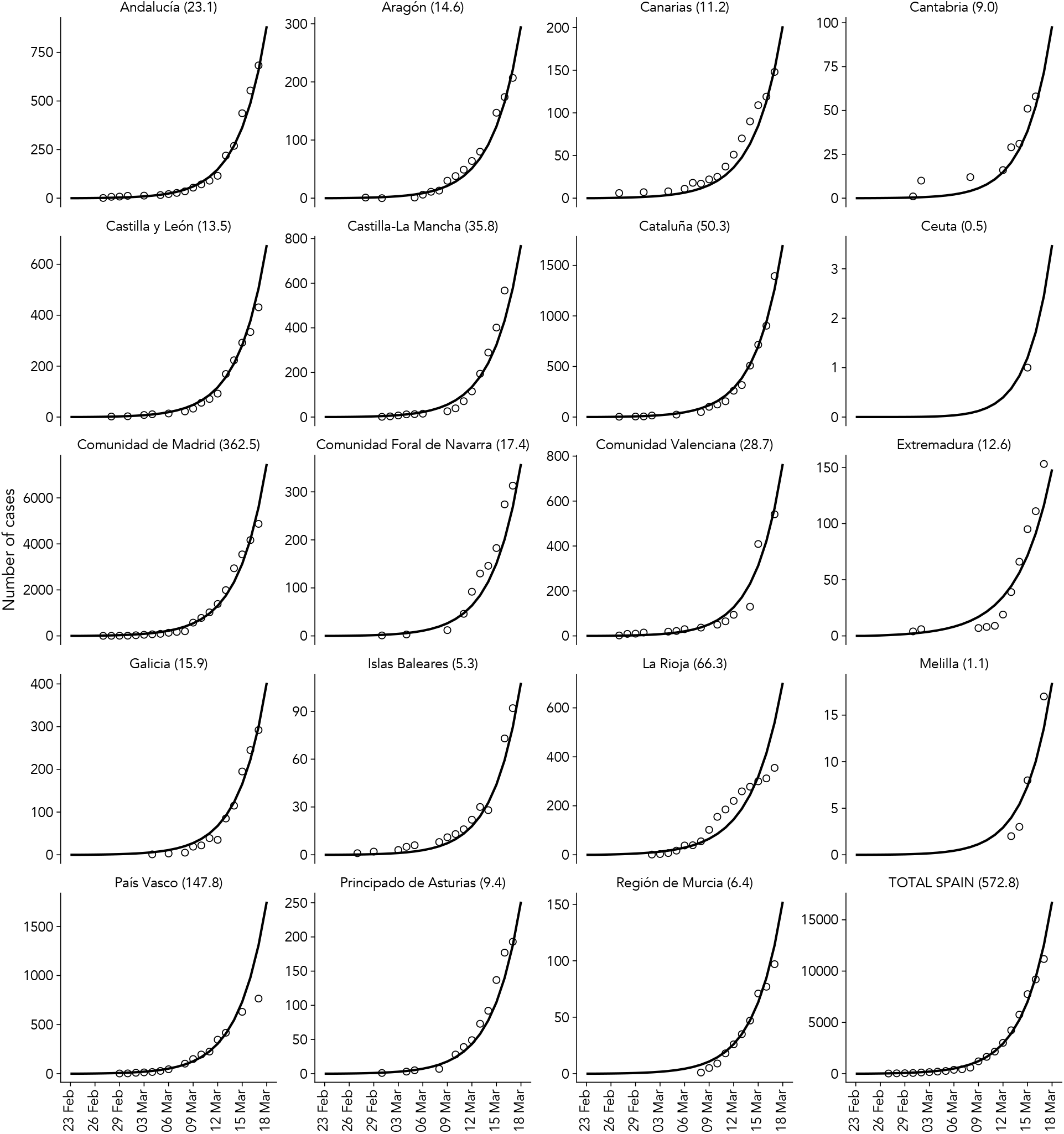
Prediction of new cases for Spain by autonomous region. Comparison of the results of the model Eqs. (S2)–(S8) for each autonomous region in Spain. The solid line is the result of the epidemic model, aggregated by ages, for the number of individuals inside compartments (H+R+D) that corresponds to the expected number of cases, and dots correspond to real cases reported. The number appearing next to the region name corresponds to the Mean Absolute Error (MAE) between the model prediction and the total number of cases.

**Supplementary Figure 3:**
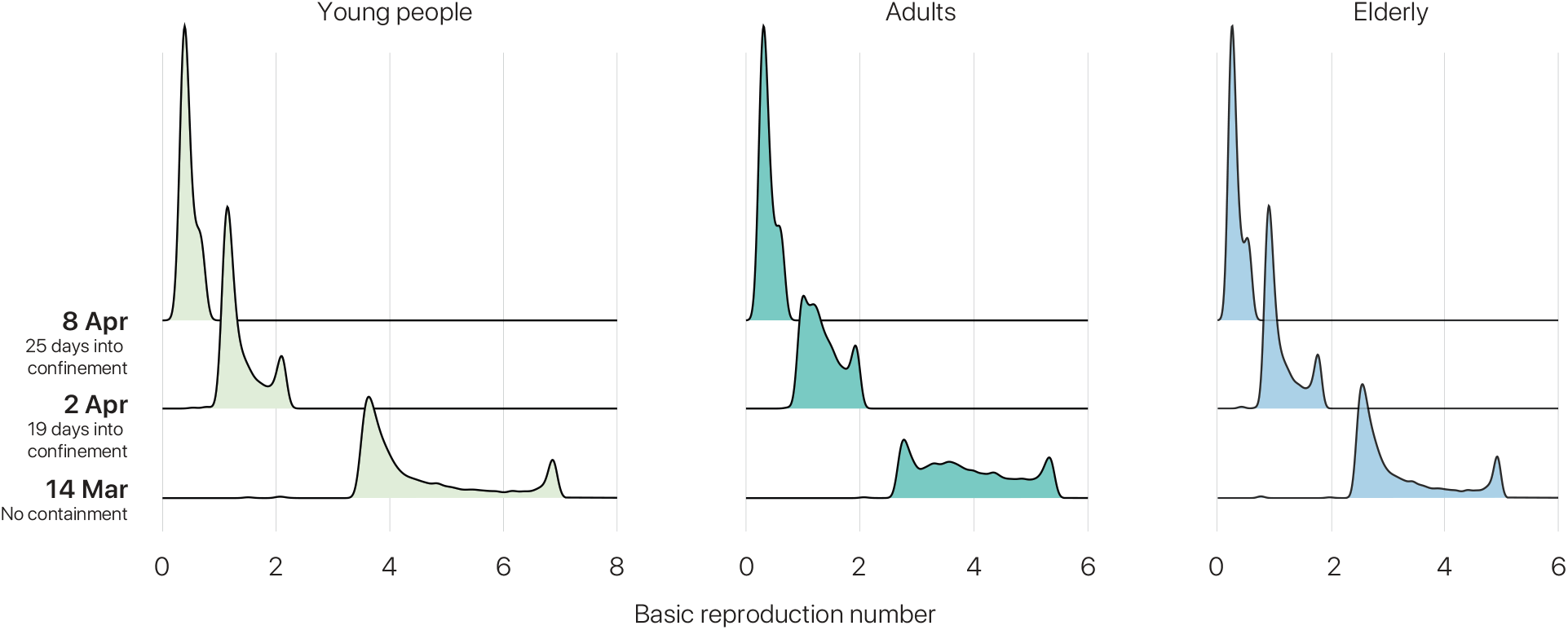
Effective reproductive number for age strata. Distribution of 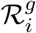 at three different stages of the epidemic spreading in Spain, according to our model: free spreading without containment (March 14, 2020); 10 days after the implementation of partial confinement, showing the initial effects of containment (March 24, 2020); and 21 days after the implementation of partial confinement (April 4, 2020).

**Supplementary Figure 4:**
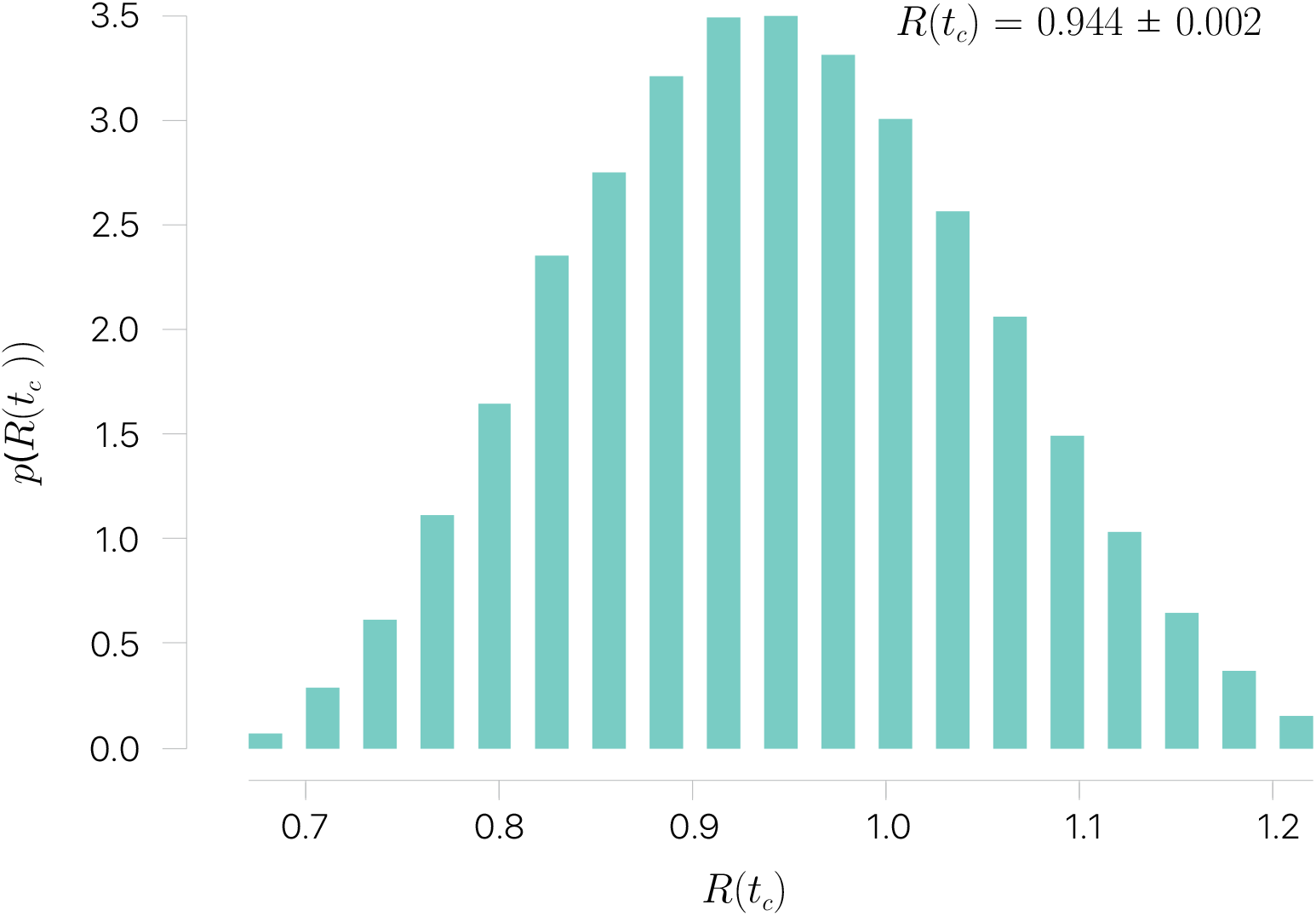
Sensitivity analysis. Distribution of effective reproduction number ℛ values while exploring the parameters space by tuning (*σ, α, ν*) as described in the text.

**Supplementary Table 1:**
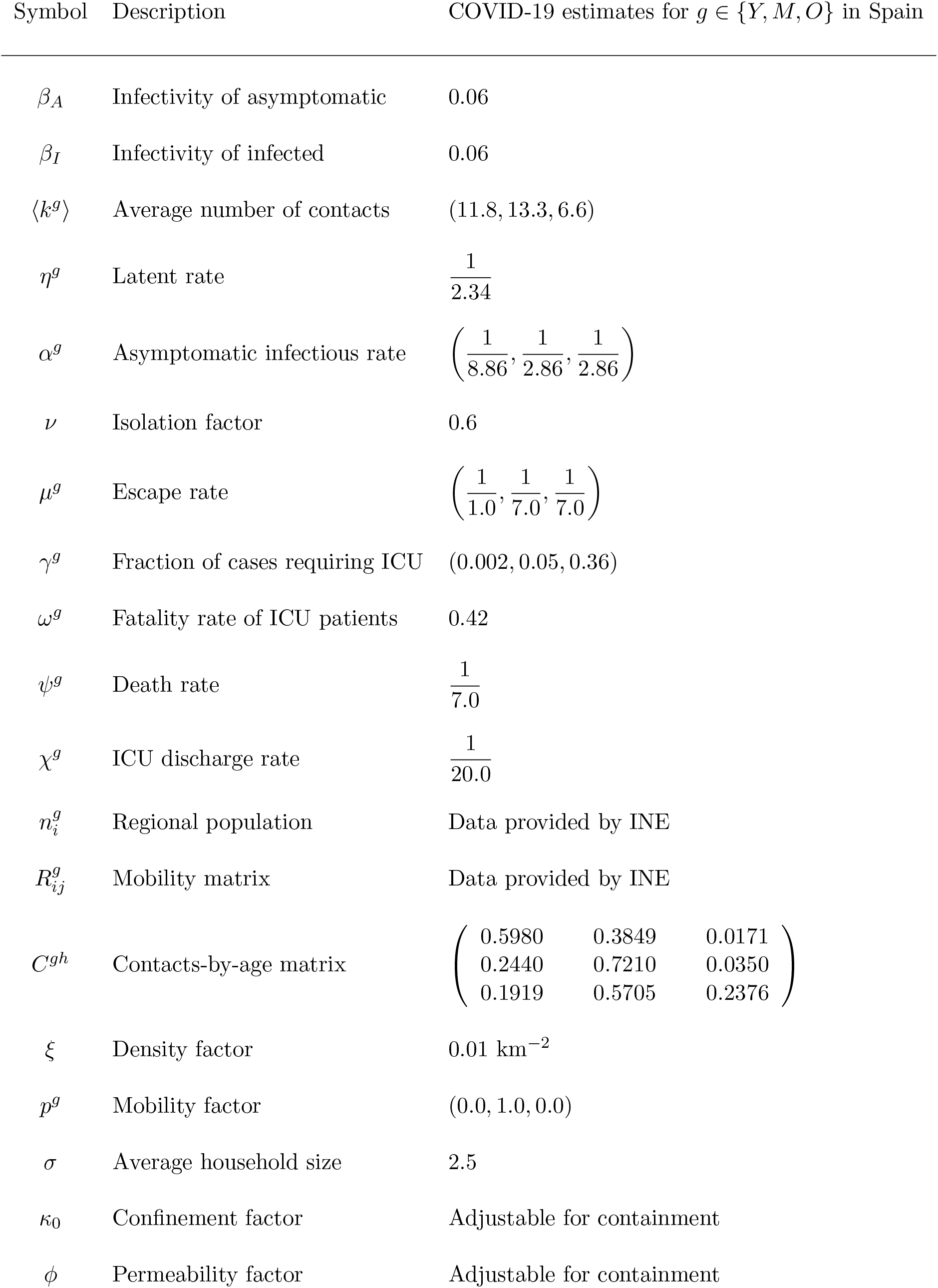
Parameters of the model and their estimations for COVID-19. See Supplementary Note 6 for a detailed explanation.

| Symbol | Description | COVID-19 estimates for $g \in \{Y, M, O\}$ in Spain |
| --- | --- | --- |
| $\beta_A$ | Infectivity of asymptomatic | 0.06 |
| $\beta_I$ | Infectivity of infected | 0.06 |
| $\langle k^g \rangle$ | Average number of contacts | (11.8, 13.3, 6.6) |
| $\eta^g$ | Latent rate | $\frac{1}{2.34}$ |
| $\alpha^g$ | Asymptomatic infectious rate | $\left( \frac{1}{8.86}, \frac{1}{2.86}, \frac{1}{2.86} \right)$ |
| $\nu$ | Isolation factor | 0.6 |
| $\mu^g$ | Escape rate | $\left( \frac{1}{1.0}, \frac{1}{7.0}, \frac{1}{7.0} \right)$ |
| $\gamma^g$ | Fraction of cases requiring ICU | (0.002, 0.05, 0.36) |
| $\omega^g$ | Fatality rate of ICU patients | 0.42 |
| $\psi^g$ | Death rate | $\frac{1}{7.0}$ |
| $\chi^g$ | ICU discharge rate | $\frac{1}{20.0}$ |
| $n_i^g$ | Regional population | Data provided by INE |
| $R_{ij}^g$ | Mobility matrix | Data provided by INE |
| $C^{gh}$ | Contacts-by-age matrix | $\begin{pmatrix} 0.5980 & 0.3849 & 0.0171 \\ 0.2440 & 0.7210 & 0.0350 \\ 0.1919 & 0.5705 & 0.2376 \end{pmatrix}$ |
| $\xi$ | Density factor | 0.01 km<sup>-2</sup> |
| $p^g$ | Mobility factor | (0.0, 1.0, 0.0) |
| $\sigma$ | Average household size | 2.5 |
| $\kappa_0$ | Confinement factor | Adjustable for containment |
| $\phi$ | Permeability factor | Adjustable for containment |

#### Supplementary Note 1. Epidemic spreading model

We propose a tailored model for the epidemic spread of COVID-19. We use a previous framework for the study of epidemics in structured metapopulations, with heterogeneous agents, subjected to recurrent mobility patterns [7, 17, 10, 18].To understand the geographical diffusion of the disease, as a result of human-human interactions in small geographical patches, one has to combine the contagion process with the long-range disease propagation due to human mobility across different spatial scales. For the case of epidemic modeling, the metapopulation scenario is as follows. A population is distributed in a set of patches, being the size (number of individuals) of each patch in principle different. The individuals within each patch are well-mixed, *i*.*e*., pathogens can be transmitted from an infected host to any of the healthy agents placed in the same patch with the same probability. The second aspect of our metapopulation model concerns the mobility of agents. Each host is allowed to change its current location and occupy another patch, thus fostering the spread of pathogens at the system level. Mobility of agents between different patches is usually represented in terms of a network where nodes are locations while a link between two patches represents the possibility of moving between them.

We introduce a set of modifications to the standard metapopulation model to account for the different states relevant for the description of COVID-19, and also to substitute the well-mixing with a more realistic set of contacts. Another key point is the introduction of a differentiation of the course of the epidemics that depends on the demographic ages of the population. This differentiation is very relevant in light of the observation of a scarcely set of infected individuals at ages (< 25), and also because of the severe situations reported for people at older ages (> 65). Our model is composed of the following epidemiological compartments: susceptible (S), exposed (E), asymptomatic infectious (A), infected (I), hospitalized to ICU (H), dead (D), and recovered (R). Additionally, we divide the individuals in *N*_*G*_ age strata, and suppose the geographical area is divided in *N* regions or patches. Although we present the model in general form, its application to COVID-19 only makes use of the three age groups mentioned above (*N*_*G*_ = 3): young people (Y), with age up to 25; adults (M), with age between 26 and 65; and elderly people (O), with age larger than 65.

Let us suppose we have a population of *N* individuals distributed in *N*_*P*_ regions, with *n*_*i*_ individuals residing in region *i*. We also consider that individuals belong to one of *N*_*G*_ different age strata, in such a way that 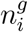 individuals of age group *g* live in region *i*. Thus,

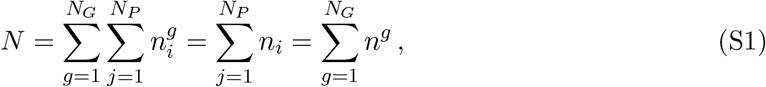

where *n*^*g*^ is the total population of group age *g*.

Our system is completely characterized with the fraction of individuals in state *m*, where *m* ∈{*S, E, A, I, H, D, R* }, for each age stratum *g*, and associated with each patch *i*, that we denote in the following as 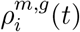. The temporal evolution of these quantities is given by:

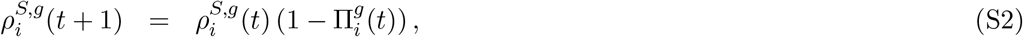

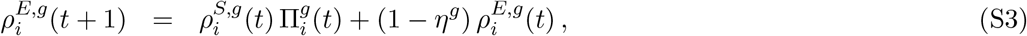

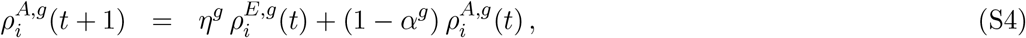

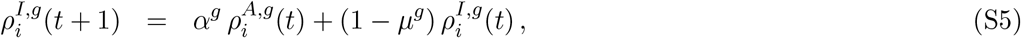

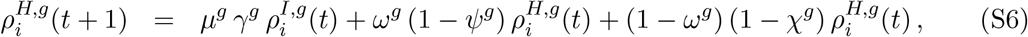

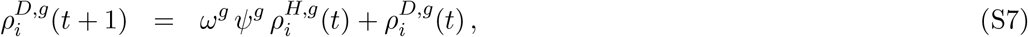

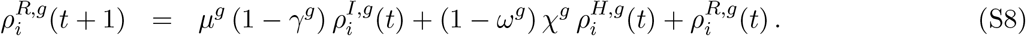

These equations correspond to a discrete-time dynamics, in which each time-step represents a day. They are built upon previous work on Microscopic Markov-Chain Approach (MMCA) modelization of epidemic spreading dynamics [6], but which has also been applied to other types of processes, *e*.*g*., information spreading and traffic congestion [8, 9, 16].

The rationale of the model is the following. Susceptible individuals get infected by contacts *I* with asymptomatic and infected agents, with a probability 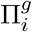, becoming exposed. Exposed individuals turn into asymptomatic at a certain rate *η*^*g*^, which in turn become infected at a rate *α*^*g*^. Once infected, two paths emerge, which are reached at an escape rate *µ*^*g*^. The first option is requiring hospitalization in an ICU, with a certain probability *γ*^*g*^; otherwise, the individuals become recovered. While being at ICU, individuals have a death probability *ω*^*g*^, which is reached at a rate *ψ*^*g*^. Finally, ICUs discharge at a rate *χ*^*g*^, leading to the recovered compartment. See in Supplementary Figure **1** a sketch of the compartmental epidemic model with all the transitions, and Supplementary Table **1** for a list of the parameters and their values to simulate the spreading of COVID-19 in Spain, which will be discussed in Supplementary Note 6.

The value of 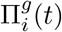 encodes the probability that a susceptible agent belonging to age group *g* and patch *i* contracts the disease. Under the model assumptions, this probability is given by:

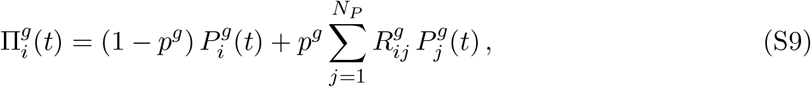

where *p*^*g*^ denotes the degree of mobility of individuals within age group 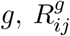 is the mobility matrix (fraction of individuals of group age *g* that choose destination *j* while living in region *i*), and 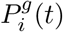 denotes the probability that those agents get infected by the pathogen inside patch *i*. This way, the first term in the r.h.s. of Eq. (S9) denotes the probability of contracting the disease inside the residence patch, whereas the second term contains those contagions taking place in any of the neighboring areas. Furthermore, we assume that the number of contacts increases with the density of each area according to a monotonously increasing function *f* . Finally, we introduce an age-specific contact matrix, *C*, whose elements *C*^*gh*^ define the fraction of contacts that individuals of age group *g* perform with individuals belonging to age group *h*. With the above definitions, 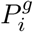 reads

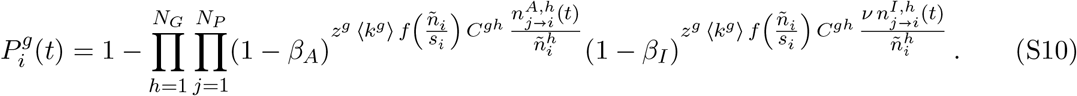

Parameters *β*_*A*_ and *β*_*I*_ correspond to the infection probabilities for contacts of a susceptible individual with asymptomatic and symptomatic infected individuals, respectively. The exponents represent the number of contacts made by an agent of age group *g* in patch *i* with infectious individuals —compartments *A* and *I*, respectively — of age group *h* residing at patch *j*. Accordingly, the double product expresses the probability for an individual belonging to age group *g* not being infected while staying in patch *i*. Finally, parameter *ν* is introduced to account for the self-isolation of symptomatic infected individuals, either because of awareness or health state, which effectively reduces their appearance in the patches.

The term *z*^*g*^⟨*k*^*g*^⟩*f* (*ñ*_*i*_*/s*_*i*_) in Eq. (S10) represents the overall number of contacts (infectious or non infectious), which increases with the density of patch *i* following function *f*, where *s*_*i*_ is the area of patch *i* measured in km^2^, and also accounts for the normalization factor *z*^*g*^, which is calculated as:

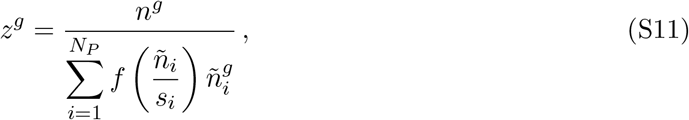

where the effective population at patch *i*, i.e., the number of people present at patch *i* when commuting takes place, is given by

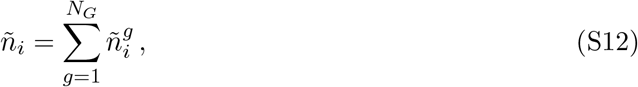

which is distributed in age groups of size

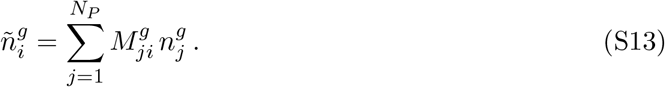

For convenience, we have defined the effective mobility matrices 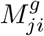,

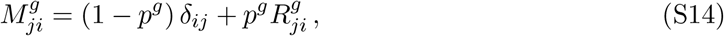

that take into account both the degree of mobility of the population, and the transition probabilities to the neighboring patches.

From now on, we will use the tilde to indicate variables measured while the commuting is active, to distinguish them from variables when all the population is in its residence patch.

The function *f*(*x*) governing the influence of population density has been selected, following [11], as:

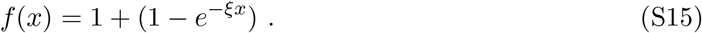

The last term of the exponents in Eq. (S10) contains the probability that these contacts are contagious, which is proportional to 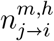, the expected number of individuals of age group *h* in the given infectious state *m* (either *A* or *I*) which have moved from region *k* to region *i*:

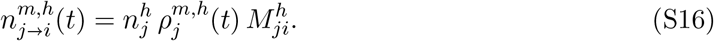

The discrete time nature of this model allows for an easy computation of the time evolution of all the relevant variables, providing information at the regional level. See Supplementary Note 7 for the details of its application to the COVID-19 outbreak in Spain. Additionally, the model is amenable for analytical inspection, which has allowed us to find the epidemic threshold, see Supplementary Note 3.

#### Supplementary Note 2. Modeling containment measures

Here we assess the performance of different containment measures to reduce the impact of COVID-19 using the mathematical model. To incorporate containment policies in our formalism, we consider that a given fraction of the adult population *κ*_0_ is isolated at home whereas both young and elderly people are assume to stay at home . In this sense, let us remark that parameter *κ*_0_ allows us to tune the strength of those containment mesaures proposed to contain COVID-19. In this sense, *κ*_0_ = 1 reflects a total lockdown of the population which isolates the households from each other, thus constraining the transmission dynamics at the level of household rather than municipality. From the former assumptions, we compute the average number of contacts of agents belonging to each group *g* ∈ {*Y, M, O*} as

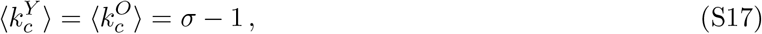

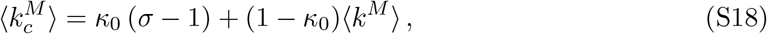

where *σ* encodes the average household size.

In this scenario, a relevant indicator to quantify the efficiency of the policy is the probability of one individual living in a household, inside a given municipality *i*, without any infected individual. Assuming that containment is implemented at time *t*_*c*_, this quantity, denoted in the following as *CH* _*i*_(*t*_*c*_), is given by

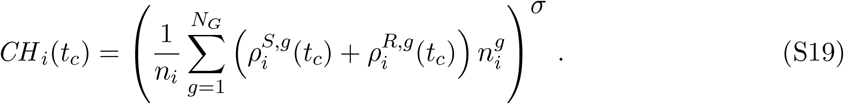

This way, Eqs. (S17) and (S18) become time-dependent:

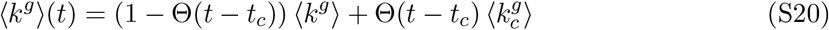

where Θ(*x*) is the Heaviside function, that is 1 if *x* ≥ 0 and 0 otherwise. Accordingly, the mobility parameters *p*^*g*^ change as

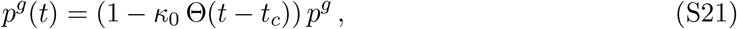

which make 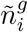 and *z*^*g*^ also dependent on time, see Eqs. (S11)–(S13). Unfortunately, complete isolation of households is impossible, thus we introduce a permeability factor *ϕ* that reduces the effective isolation.

This containment strategy is introduced in the dynamical Eqs. (S2)–(S7) by modifying Eqs. (S2) and (S3) for the time after *t*_*c*_:

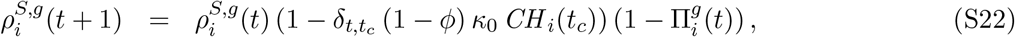

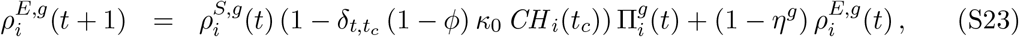

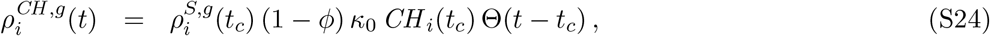

where we have added a new compartment *CH* to hold the individuals under household isolation after applying containment *κ*_0_, and *δ*_*a*,*b*_ is the Kronecker function, which is 1 if *a* = *b* and 0 otherwise. Containment also affects the average number of contacts, thus we must also update Eq. (S10):

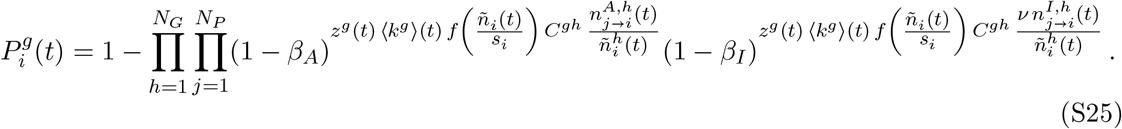

In summary, we have introduced a containment strategy characterized by the isolation of a certain fraction of the population, which in turn reduces the average number of contacts, the mobility and, as a result, the probability of becoming infected, thus reducing the overall prevalence of the disease. The containment is parameterized with the containment factor *κ*_0_, and the permeability factor *ϕ*.

#### Supplementary Note 3. Calculation of ℛ_0_ using the next generation matrix

The model is amenable for analytical calculations. We calculate the basic reproduction number ℛ_0_ using the next generation matrix (NGM) approach [4]. Accordingly, we need to analyze the stability of the disease free equilibrium. We do so by making a first order expansion of the model equations for small values (*ϵ*) of the non-susceptible states *m*: 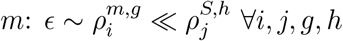 and 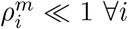, where *m* ∈ {*E, A, I, H, D, R*}. The expansion allows us to transform our discrete time Markov Chain into a system of continuous time differential equations. We start by expanding the infection probabilities 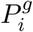:

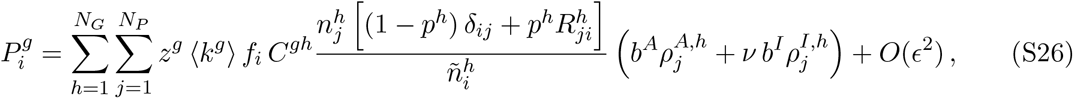

where we have defined

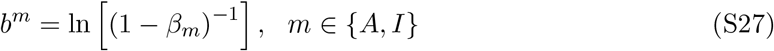

and

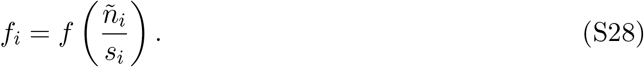

We then insert the above expression into 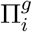, leading to:

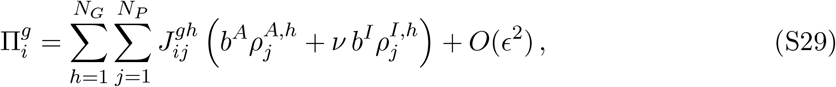

where it is convenient to separate tensor *J* if four terms:

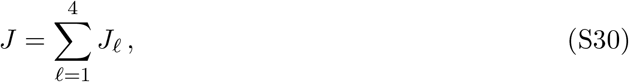

With

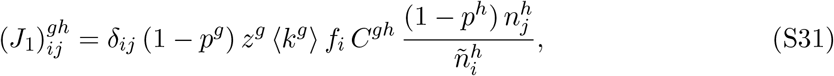

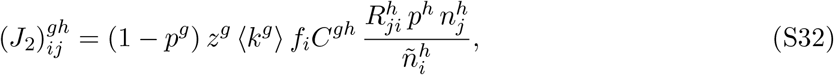

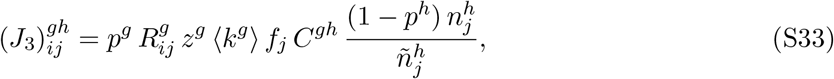

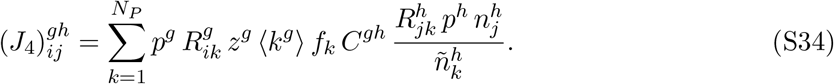

These tensors encode the four different ways in which the epidemic interactions may take place: individuals belonging to the same patch *i* = *j* and not moving (*J*_1_); interaction in the patch of *i* with individuals coming from patch *j* (*J*_2_); interaction in the patch of *j* with individuals coming from patch *i* (*J*_3_); and individuals from *i* and *j* interacting at any other patch *k* (*J*_4_).

In the next generation matrix framework, we only need to consider the epidemic compartments. Additionally, the framework requires to work in absolute numbers of infected individuals. Accordingly, we define the absolute number of individuals in the respective compartments as

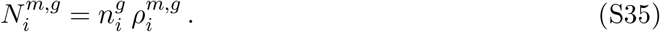

Making use of the above definitions, the corresponding differential equations take the form:

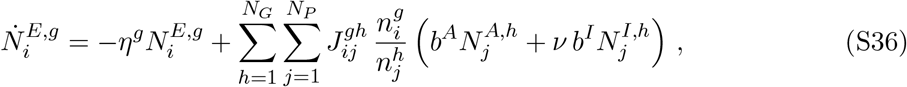

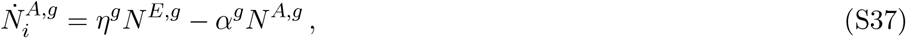

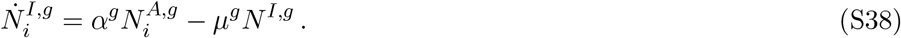

Defining the vector *N*^*g*^ =(*N*^*E*,*g*^, *N*^*A*,*g*^, *N*^*I*,*g*^)*T*, the above system of differential equations can be rewritten as:

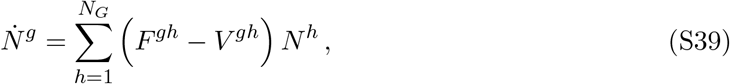

where we have defined

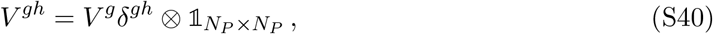

with,

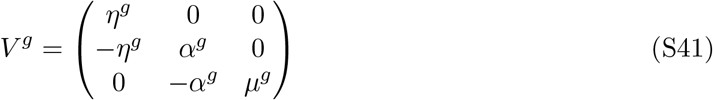

and

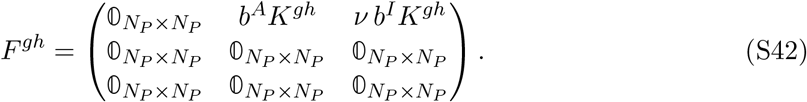

In the above tensor we have defined

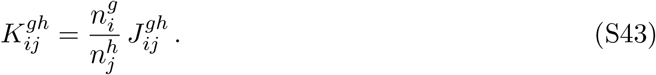

With the above differential equation, the basic reproduction number is given by:

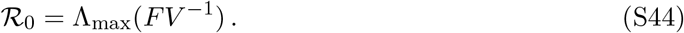

We can calculate the inverse of the tensor *V* as

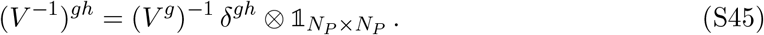

The inverse of the matrix *V* ^*g*^ is given by:

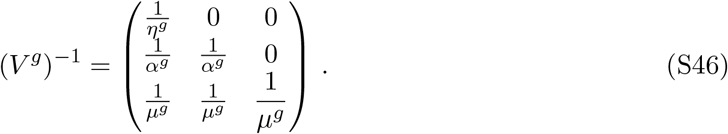

Accordingly, we have:

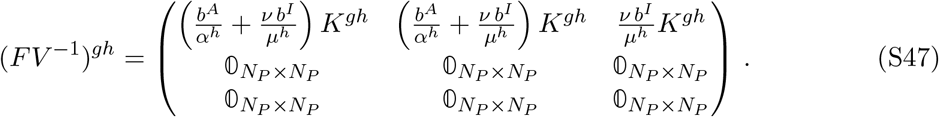

As we are looking for the eigenvectors of the tensor *FV* ^−1^, we note that their components associated to the compartments *A* and *I* —rows 2 and 3— must be zero, since the associated rows in the above matrix are zero. To be more precise, we have 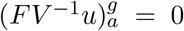 for *a* = 2*N*_*P*_ +1, … 3*N*_*P*_, which are the elements associated to the compartments *A* and *I*. Accordingly, we can restrict the above matrix only to the vector space associated to the compartment *E* and the eigenvalues will be equivalent, which gives us the basic reproduction number ℛ_0_:

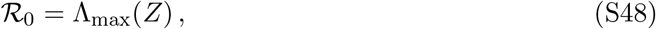

Where

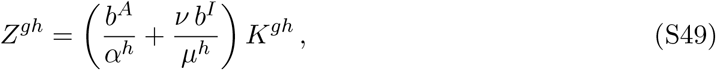

and Λ_max_(*Z*) denotes the spectral radius of tensor *Z*.

#### Supplementary Note 4. Effective reproduction number and critical containment

The effective reproduction number ℛ is given by

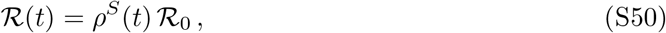

where *ρ*^*S*^(*t*) is the fraction of reachable susceptible individuals at time *t* and ℛ_0_ is the basic reproduction number [3]. In our case, the containment has an effect on both terms. First, *κ*_0_ reduces the pool of individuals susceptible of contracting the disease, i.e. it reduces the term *ρ*^*S*^. And, second, *κ*_0_ reduces the average degree of the individuals and thus it reduces ℛ_0_. Eq. (S50) allows us to accurately determine the effective reproduction number and capture the impact of human mobility and containment measures on the spread of the disease.

Computing Eq. (S50) by means of the NGM approach involves the calculation of the spectral radius of tensor *Z*, which hinders an understanding of the roots behind the transition from flattening to bending the epidemic curve triggered by an efficient confinement, as illustrated in the main text. In order to have an analytical estimation of the critical value of the needed confinement to observe this transition, 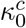, let us consider a single well mixed population. In this case, we have

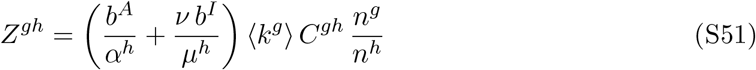

and

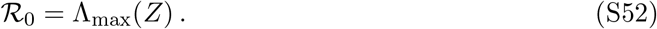

If we remove the age strata, the basic reproduction number is reduced to

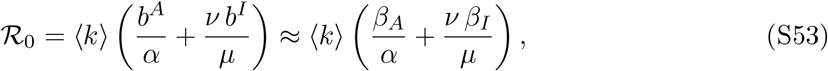

where we have approximated

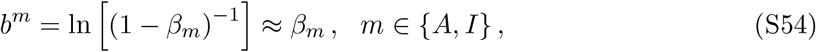

which is valid for low values of the infection probabilities *β*_*m*_. The same result for ℛ_0_, Eq. (S53), is obtained if all age groups share the values of population *n*^*g*^, average number of contacts ⟨*k*^*g*^⟩, asymptomatic infectious rate ⟨*α*^*g*^⟩, and escape rate ⟨*µ*^*g*^⟩, since Λ_max_(*C*) = 1 due to the contact-by-age matrix *C* being a stochastic matrix.

Accordingly, after introducing containment, we get for the effective reproduction number:

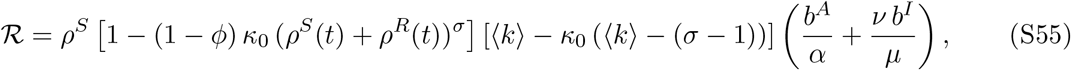

where the temporal dependence has been omitted. The parameter *ϕ* encodes the permeability (household isolation level), ranging from *ϕ* = 0 for which households are perfectly isolated, to *ϕ* = 1 which recovers the well-mixed scenario before the intervention, but now with a lower average number of contacts.

In order to bend the epidemic curve and not to flatten it we impose the condition ℛ ≤ 1. Accordingly, the critical containment value 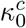 must fulfill the equation:

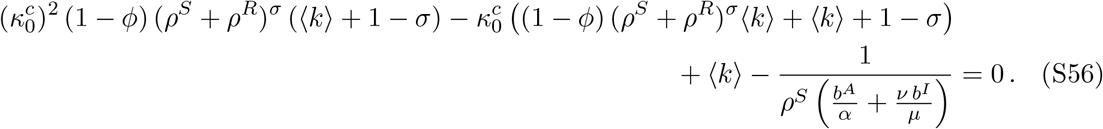

Defining the following coefficients:

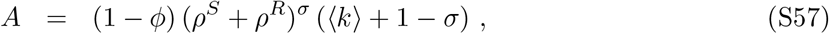

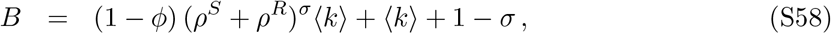

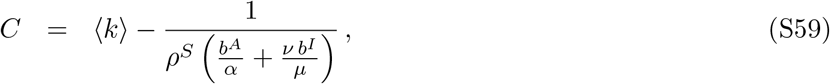

we can write Eq. (S56) as

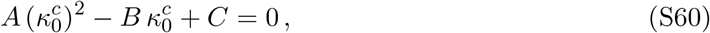

whose solution is given by:

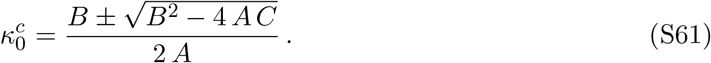

Note that only the physically meaningful solution, 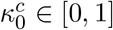, must be retained.

#### Supplementary Note 5. Dynamical approximation of the effective reproduction number

A naive approach to computing the effective reproduction number ℛ is considering the mean field scenario, in which an infected subject *i* contacts ⟨*k*⟩ individuals each time step. Assuming an infection probability *β*, the expected number of individuals infected by *i* at each time step is *ρ*_*S*_ ⟨*k*⟩*β*, where *ρ*_*S*_ is the fraction of susceptible individuals on the population. Taking into account the duration of the infectious period *τ*, we can estimate how many individuals have been infected by subject *i* over time as: ℛ = *ρ*_*S*_ ⟨*k*⟩*βτ* .

We can extend this approach to include both the temporal dependence and the heterogeneity of contacts induced by daily commuting patterns, making use of all the probabilities provided by our model equations. Moreover, here we aim at studying the temporal evolution of the effective reproduction number beyond the early stage of the disease. In this sense, we define the reproduction number of patch *i* and age group *g* as the number of infections observed if we seed an infected individual in the aforementioned patch and age group [14]. This involves considering that this infected individual, with residence in patch *i*, may commute to patch *j*, where it will be able to contact and infect susceptible individuals coming from any other patch *k*.

First, we calculate the expected number of susceptible individuals of age group *h* which have moved from region *k* to region *j* as:

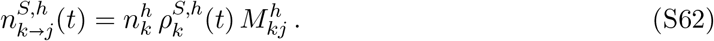

The fraction of susceptible individuals of each age group *g* in patch *j* at time *t* is expressed by:

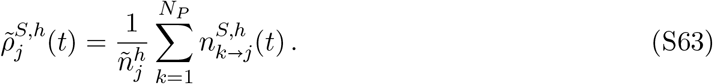

Next, we compute the number of susceptible contacts made by an individual of age group *g* and patch *i*, which can be expressed as:

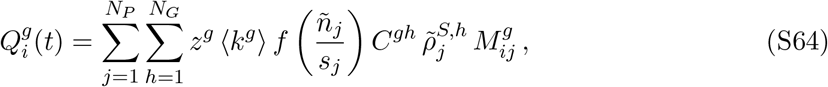

Using this expression, and assuming that we have an individual from patch *i* and age group *g* that has become infectious, we can compute the effective reproduction number for each patch and age group as:

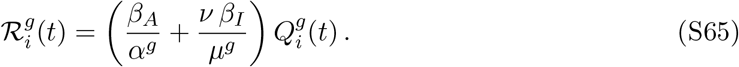

For estimating the global effective reproduction number, we then make use of a weighted average taking into account the distribution of the population across patches and age groups:

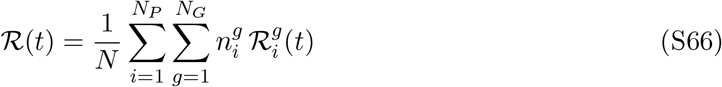

In Eq. (S65), we assume that the fraction of susceptible individuals as well as the average number of contacts is constant during the infectious period. However, the fraction of susceptible individuals obviously changes during that time. More importantly, as containment measures are put in place, the number of contacts varies as well. To account for the temporal variability of these quantities, we calculate the contributions to 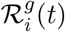 of the infections produced since the individual became asymptomatically infected at time *t*. Since the infectious period is divided in two phases, one asymptomatic with infectious rate *α*^*g*^, and another symptomatic with infectious rate *µ*^*g*^, what we have is a coupled Poisson process. The evolution of the probabilities to be asymptomatic, *ζ*^*A*,*g*^, or symptomatic, *ζ*^*I*,*g*^, are given by:

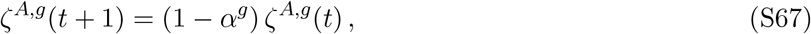

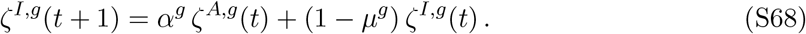

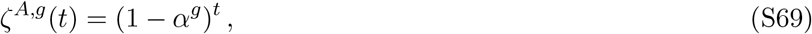

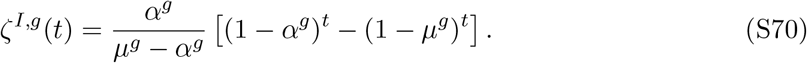

Incorporating these probabilities into Eq. (S65) we obtain

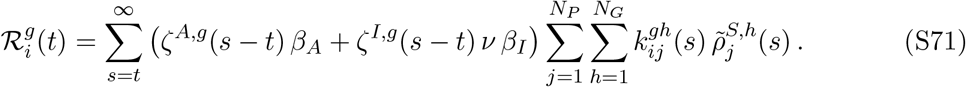

#### Supplementary Note 6. Parameters for the modelization of the spreading of COVID-19

In this subsection, we detail our parameters choice to study the current epidemic outbreak in Spain. Regarding epidemiological parameters, the incubation period has been reported to be *η*^−1^ + *α*^−1^ = 5.2 days [12] in average which, in our formalism, must be distributed into the exposed and asymptomatic compartments. In principle, if one does not expect asymptomatic transmissions, most of this time should be spent inside the exposed compartment, thus being the asymptomatic infectious compartment totally irrelevant for disease spreading. However, along the line of several recent works [13, 15, 5] we have found that the unfolding of COVID-19 cannot be explained without accounting for infections from individuals not developing any symptoms previously. In particular, our best fit to reproduce the evolution of the real cases reported so far in Spain yields *α*^−1^ = 2.86 days as asymptomatic infectious period. In turn, the infectious period while being symptomatic is established as *µ*^−1^ = 7 days [1, 20], except for the young strata, for which we have reduced it to 1 day, assigning the remaining 6 days as asymptomatic; this is due to the reported mild symptoms in young individuals, which may become inadvertent [2]. Furthermore, we have included an isolation factor *ν* = 0.6 which reduces the infectiousness of symptomatic agents as a result of their self-isolation. We fix the fatality rate *ω* = 42% of ICU patients by studying historical records of dead individuals as a function of those requiring intensive care. In turn, we estimate the period from ICU admission to death as *ψ*^−1^ = 7 days [19] and the stay in ICU for those overcoming the disease as *χ*^−1^ = 20 days [1].

#### Supplementary Note 7. Calibration and validation of the model for the evolution of COVID-19 in Spain

Equations (S2)–(S8) enable to monitor the spatio-temporal propagation of COVID-19 across Spain. To check the validity of our formalism, we aggregate the number of cases predicted for each municipality at the level of autonomous regions (*comunidades autónomas*), which is a first-level political and administrative division, and compare them with the number of cases daily reported by the Spanish Health Ministry. In this sense, we compute the number of cases predicted for each municipality *i* at each time step *t* as:

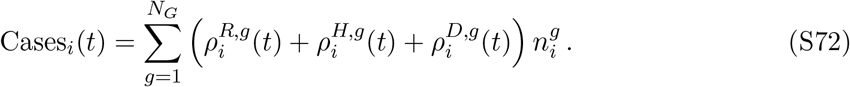

As our model is designed to predict the emergence of autochthonous cases triggered by local contagions and commuting patterns, those imported infected individuals corresponding to the first reported cases in Spain are initially plugged into our model as asymptomatic infectious agents. In addition, small infectious seeds should be also placed in those areas where anomalous outbreaks have occurred due to singular events such as one funeral in Vitoria leading to more than 60 contagions. Overall, the total number of infectious seeds is 47 individuals which represents 0.2 % of the number of cases reported by March 20, 2020.

Supplementary Figure **2** shows that our model is able to accurately predict not only the overall evolution of the total number of cases at the national scale but also their spatial distribution across the different autonomous regions. Moreover, the most typical trend observed so far is an exponential growth of the number of cases, thus clearly suggesting that the disease was spreading freely in most of the territories when the model was calibrated. Note, however, that there are some exceptions such as La Rioja or País Vasco in which some strong policies targeting the most affected areas were previously promoted to slow down COVID-19 propagation.

We also show in Supplementary Figure **3** the distribution of 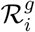 at three different stages of the epidemic spreading in Spain, according to our model: free spreading without containment (March 14, 2020); 10 days after the implementation of partial confinement, showing the initial effects of containment (March 24, 2020); and 21 days after the implementation of partial confinement (April 4, 2020).

#### Supplementary Note 8. Sensitivity Analysis

We have performed a sensitivity analysis to check the robustness of the critical value of the effective reproduction number ℛ = 1, identifying the transition from supercritical (flattening) regime to the subcritical (bending) one when applying confinement. For this purpose, we report here the results obtained by tuning some of the parameters of the model under the same assumptions and show that the obtained results are consistent with the results reported in the main text of the manuscript and do not alter the conclusions of our study.

In particular, we fix the permeability parameter to *ϕ* = 0.20 and the confinement to *κ*_0_ = 0.66, and sample a region of the parameters space by varying the average household size, *σ* ∈ [2, 3], the asymptomatic infectious rate of each strata *g, α*^*g*^ ∈ [0.25, 0.5], and the self-isolation parameter of symptomatic infected individuals, *ν* ∈ [0.5, 0.7].

Results of this sensitivity analysis are depicted in Figure **4**, where we show that the effective reproduction number remains almost unaltered while exploring different points of the parameters space.

## Notes

### Competing Interest Statement

The authors have declared no competing interest.

### Funding Statement

Spanish MINECO (grant PGC2018-094754-B-C21, FIS2015-71582-C2 and FIS2017-87519-P),
Generalitat de Catalunya (grant No. 2017SGR-896),
Universitat Rovira i Virgili (grant No. 2018PFR-URV-B2-41)
James S. McDonnell Foundation grant #220020325
Departamento de Industria de Innovación del Gobierno de Aragón y Fondo Social Europeo (FENOL group E-19)
Ministerio de Ciencia, Innovación y Universidades (program JdC)
Marie Sklodowska-Curie grant agreement No. 713679
Coordenacao de Aperfeicoamento de Pessoal de Nıvel Superior, Brasil
(CAPES), Finance Code 001

